# Uncovering the Biological Toll of Neighborhood Physical Disorder: Links to Metabolic and Inflammatory Biomarkers in Older Adults

**DOI:** 10.1101/2024.12.23.24319571

**Authors:** Jiao Yu, Thomas K.M. Cudjoe, Walter S. Mathis, Xi Chen

## Abstract

**Background:** Neighborhood physical disorder has been linked to adverse health outcomes, yet longitudinal assessments of its relationship with metabolic and inflammatory biomarkers in older adults remain limited. This study examined the association between patterns of neighborhood physical disorder exposure and biomarkers among older adults.

**Methods:** We included community-dwelling Medicare beneficiaries with 2017 biomarker data from the National Health and Aging Trends Study (n = 4,558). Neighborhood physical disorder from 2011 to 2016 was assessed using interviewer reports of neighborhood characteristics. Latent class analysis was employed to identify longitudinal patterns of exposure. Inverse probability weighted linear regression models were used to examine associations between physical disorder patterns and five biomarkers, including body mass index (BMI), waist circumference, hemoglobin A1C (HbA1c), high-sensitivity C-reactive protein (hsCRP), and interleukin-6 (IL-6).

**Results:** Four classes of neighborhood physical disorder emerged: stable low exposure (85%), increased exposure (4%), decreased exposure (8%), and stable high exposure (3%). Regression findings indicate that residing in neighborhoods with stable high exposure was significantly associated with higher levels of BMI (*b = 0.06, p<0.05*), HbA1c (*b* = 0.09, *p<0.05*), hsCRP (*b = 0.21, p < 0.05*), and IL-6 (*b = 0.22, p < 0.05*), compared to those with stable low exposure. Older adults with increased exposure and decreased exposure also exhibited elevated risks in multiple metabolic and inflammation biomarkers.

**Conclusions:** Persistent exposure to neighborhood physical disorder is associated with higher levels of metabolic and inflammatory biomarkers, underscoring the need for targeted clinical screening and neighborhood initiatives to promote healthy aging in place.

---

The neighborhood health effects have been well established. Neighborhood physical disorder, ^1^ characterized by visible signs of decay and deterioration such as broken windows, littered streets, and vacant buildings, has emerged as a significant risk factor for physical dysregulation, functional limitations, and various health conditions in old age. ^2–5^ Recent studies delving into the physiologic pathways between neighborhood environments and health have focused on metabolic and inflammatory biomarkers, as they are considered among the most important indicators of cardiovascular diseases,^6,7^ diabetes,^8^ elevated morbidity, and mortality.^9^ For instance, neighborhood physical disorder is significantly associated with inflammatory markers, such as CRP and IL-6.^10^ Living in a deprived neighborhood is linked to higher body mass index (BMI), increased waist circumference, and elevated hemoglobin A1C (HbA1c) levels. ^11,12^ Collectively, these biomarkers may reflect distinct yet interrelated pathways through which neighborhood physical disorder elevates the biological risk of aging and increases vulnerabilities in older populations. ^13^

Among the various mechanisms proposed to explain the link between neighborhood physical disorder and adverse health outcomes, the stress response pathway may be the most relevant. ^14,15^ Living in disordered neighborhoods can be inherently stressful. Residents in such neighborhoods report greater exposure to and intensity of daily stressors and hassles. ^16,17^ Chronic exposure to neighborhood physical disorder can trigger heightened vigilance and sustained stress responses. Physiological and psychological stress activates the hypothalamic-pituitary-adrenal (HPA) axis and the sympathetic nervous system (SNS), leading to the release of pro-inflammatory cytokines like interleukin-6 (IL-6) and C-reactive protein (CRP).^8,18^ Chronic stress and the resulting low-grade inflammation are linked to metabolic dysregulation, manifesting in elevated hemoglobin A1c (HbA1c), metabolic abnormalities, and anthropometric changes. ^19–21^

Despite the well-documented associations between neighborhood physical disorder and metabolic and inflammatory biomarkers, much of the existing research relies on cross-sectional data, providing only a snapshot of these relationships at a single point in time. This cross-sectional approach not only fails to account for the cumulative exposure to neighborhood physical disorder but also aggravates the concern of reverse association. ^3,12^ Recent review articles further suggest that prolonged exposure, often spanning several years, is typically required for the effects of neighborhood risks on health to become evident. ^22,23^ This emphasizes the need for a longitudinal design to better understand the extent to which neighborhood environments are linked to health and well-being. In light of this gap, our study shifts the focus from static measurements of neighborhood physical disorder to a more dynamic perspective, considering both the duration and trajectory of exposure. We aim to identify distinct groups based on changing exposures to neighborhood physical disorder among older individuals in those contexts.

Residential mobility is selective and neighborhoods also change over time.^24^ Individuals are often non-randomly sorted into neighborhoods based on socioeconomic status, health conditions, or other individual-level characteristics. For instance, individuals of higher socioeconomic status tend to live in advantaged neighborhoods, while those with fewer resources may stay in less advantaged ones. This sorting process presents a challenge in disentangling the neighborhood effects from individual circumstances.^13,22^ To address this issue, recent research has employed inverse probability weighting (IPW) adjustments to account for selection into neighborhoods and disentangle neighborhood effects from the confounding effects.^13^ In this study, we employed a machine learning-based IPW method to balance respondent characteristics across neighborhoods and mitigate the potential confounding.

Using a nationally representative longitudinal sample of older adults from the National Health and Aging Trends Study (NHATS, 2011 - 2017), we employed latent class analysis to rigorously assess the association between neighborhood physical disorder history and metabolic and inflammatory biomarkers. Specifically, we aim to answer two research questions: (1) What are the patterns of neighborhood physical disorder experienced by these older adults over the study period? (2) What are the relationships between exposure to different patterns of neighborhood physical disorder and metabolic and inflammation biomarkers for this population-based cohort of older adults? Given the established relationship between adverse neighborhood conditions and disease risks, we hypothesize that living in neighborhoods with higher physical disorder is associated with higher levels of metabolic syndrome and inflammation as indicated by metabolic and inflammatory biomarkers.

## Study Sample

The National Health and Aging Trend Study (NHATS) is a nationally representative longitudinal cohort study of Medicare beneficiaries 65 years and older. The NHATS respondents were recruited from the Medicare enrollment database using a stratified three-stage sample design. These respondents have been followed annually since its inception in 2011. A new replenished cohort was introduced in 2015 to restore the panel to its original size. Detailed information about the NHATS study design and procedures can be found elsewhere.^25^ For this study, we utilized respondents’ information from Rounds 1 (2011) to Round 7 (2017). Key social-demographic information, such as race/ethnicity, gender, and education, was obtained upon entry. Neighborhood physical disorder was repeated measures from Round 1 to Round 6. Blood biomarkers and anthropometric markers were collected in 2017 (Round 7). All respondents who completed the 2017 interview were invited to participate in a dried blood spot (DBS) collection, except for those whose interviews were conducted by proxies. Among the eligible respondents (n = 5,266), 93% (n = 4,903) consented to blood collection. Of those who consented, 95.7% (n = 4,691) successfully provided a blood specimen. This research excluded nursing home respondents (n = 37), as neighborhood characteristics were less relevant to those older adults. We also excluded respondents with missing data on outcomes or covariates (n = 96), resulting in a final analytic sample of 4,558 older adults. Figure 1 shows the inclusion and exclusion criteria for this study.

**Figure 1.**
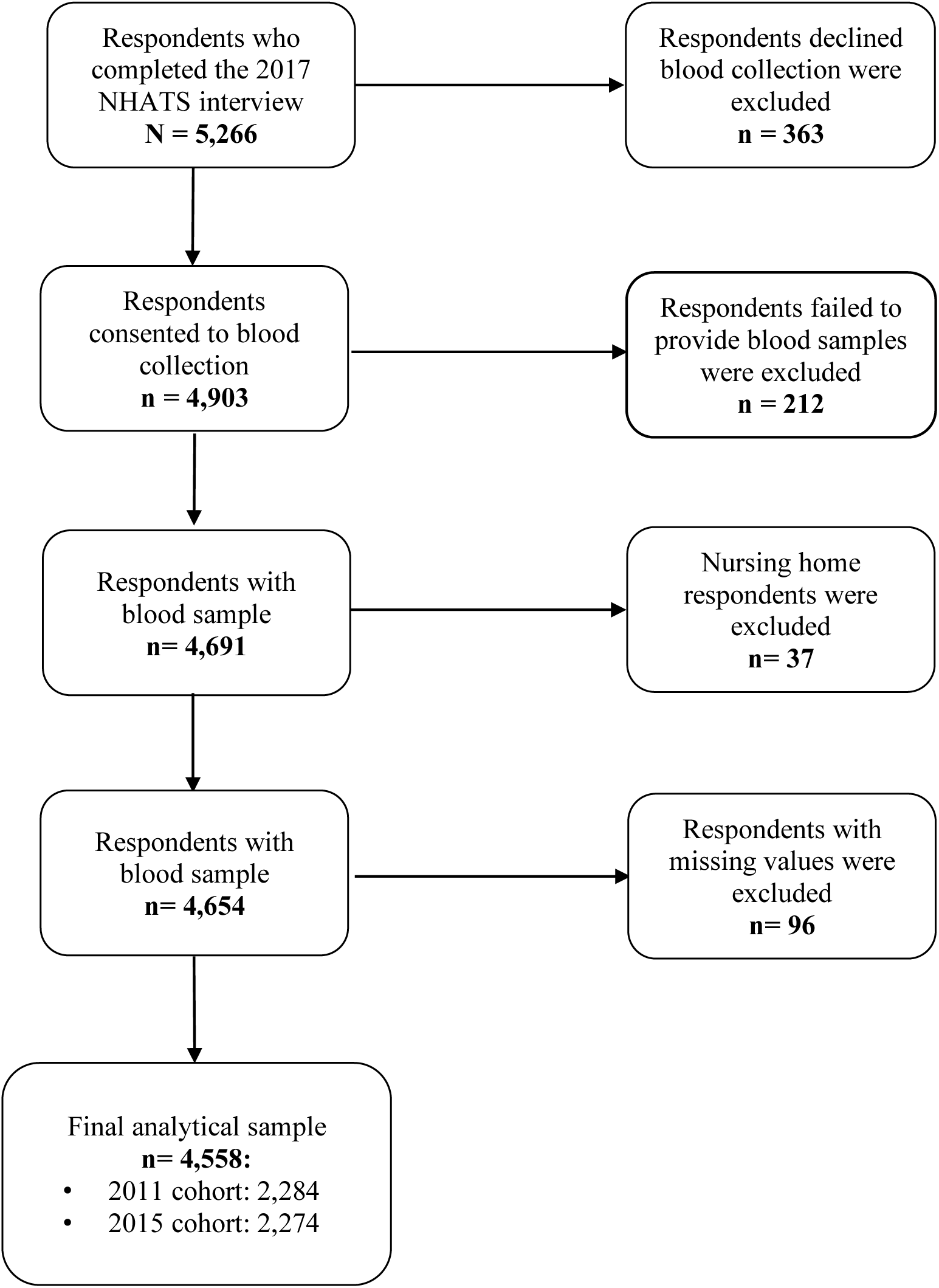
Analytic Sample Selection Process

## Measures

### Metabolic and inflammatory biomarkers

We included five biomarkers from the Round 7 DBS survey reflecting metabolism, adiposity, and inflammation. Those included hemoglobin A1C (HbA1c) in %, body mass index (BMI), waist circumference in cm, high-sensitivity CRP (hsCRP) in mg/L, and interleukin-6 (IL-6) in pg/mL. HbA1c, hsCRP, and IL-6 were assayed based on the dried blood spot. Detailed assay procedures are described elsewhere.^26^ Assay results were available as direct analyte concentrations or plasma-equivalent concentrations. We used plasma-equivalent values to facilitate clinical interpretation and comparison with other published metrics.^13^ The distributions of all biomarkers were skewed; thus, log transformations were applied prior to the analysis.

### Neighborhood physical disorder

Using data from Round 1 through Round 6, neighborhood physical disorder was measured with a 3-item environmental checklist completed by the NHATS interviewers based on their observations of the neighborhood environments around the respondents’ residences. The interviewers documented the extent to which they observed the following when standing in front of respondents’ homes/buildings: 1) litter or trash on the ground; 2) graffiti on walls; 3) vacant homes or stores. Responses were recorded on a 4-point scale: none (1), a little (2), some (3), and a lot (4). A total score was calculated for each survey round by summing these items, with higher scores indicating greater physical disorder (Cronbach’s α ranges from 0.82 to 0.98). Because the distributions of neighborhood physical disorder measures were highly skewed and signs of disorder were rare (only around 10% for each survey round), we dichotomized the scale to indicate no physical disorder (0) and the presence of any physical disorder (1) for each survey round.

### Covariates

We included a rich array of demographic information, early-life characteristics, health behaviors, geographic factors, and housing information in the IPW model to account for the unbalanced distribution of individual-level characteristics across neighborhood physical disorder groups. These covariates included gender (male or female), race/ethnicity (white, black, Hispanic, or others), age groups (65–69, 70–74, 75–79, 80–84, 85–89, or 90 and older), educational attainment (less than high school, high school graduate or GED, some college but no degree, or college degree or above), nativity status (whether born in the United States), childhood financial status (poor, average, or good), marital status (separated/divorced/widowed/never married or married/partnered), ever smoking (no or yes), experiencing financial strain (defined as any of the following: lack of money for 1) the rent/mortgage, 2) utility bills, or 3) medical/prescription bills in the past month or 4) skipping meals in the past month), home ownership (rented or not rented), urbanicity (whether reside in a metropolitan area), and U.S. regions based on the Census classification (New England, Middle Atlantic, East North Central, West North Central, South Atlantic, East South Central, West South Central, Mountain Division, or Pacific Division). All covariates were measured at the baseline (respondents’ entry round). Since health conditions likely lie on the pathway between neighborhood physical disorder and biological risks, health variables were not controlled for in these analyses.^27^

### Statistical Analysis

We conducted latent class analysis (LCA) to identify neighborhood physical disorder groups using data from Round 1 to Round 6. LCA is a finite mixture model that assumes a finite number of unobserved subgroups, or latent classes within a population based on observed categorical data. Utilizing maximum likelihood estimation, individuals are assigned to distinct latent classes according to their estimated posterior membership probabilities. This method is particularly useful to uncover underlying structures and identify heterogeneous subgroups within a population. In this study, we first conducted multiple imputation for respondents who had missing values in neighborhood physical disorder measures. Then, we tested two- to five latent class solutions to determine the most parsimonious and statistically meaningful classification. The optimal class solution was determined based on model fit indices including the Bayesian Information Criterion (BIC) and the g-squared likelihood ratio chi-square test, combined with a graphical examination to assess whether a certain number of latent classes provided a clearer theoretical interpretation of the data. Once the best model was selected, participants were classified into subgroups according to their posterior membership probabilities. We then performed a bivariate analysis to cross-tabulate class membership with respondents’ baseline characteristics.

Next, we conducted regression analyses to assess the association between the identified neighborhood physical disorder subgroups and biological risks of aging, i.e., metabolic and inflammatory biomarkers (log-transformed) as outcome variables. Observational studies examining neighborhood effects on health outcomes may be biased owing to differences in individual characteristics related to both neighborhood exposure and health outcomes or the lack of an equivalent control group.^28^ To address this issue, we employed regression analyses estimated with inverse probability weights (IPW). This approach allows us to mimic a quasi-experimental design to estimate the treatment effects of neighborhood exposure on biomarkers while accounting for differential probabilities of exposure to various patterns of neighborhood physical disorder.

The regression analysis proceeded in two steps. First, a machine learning-based generalized boosted tree model (GBM) was employed to estimate the propensity score by including a comprehensive set of baseline covariates, including age, gender, race/ethnicity, education, marital status, nativity status, childhood financial status, ever smoking, current financial strain, home ownership, urbanicity, and residential regions.^13^ The GBM approach is preferred when dealing with multiple treatment groups and is more flexible in accommodating nonlinear functional forms, interactive effects, and model misspecification.^29^ Diagnostic assessments of covariate balance were conducted utilizing the Kolmogorov-Smirnov statistic and standardized differences. Covariate balance was considered achieved if the standardized mean difference was less than 0.1 and the Kolmogorov-Smirnov statistic was non-significant. IPW were then calculated with the inverse of the propensity scores and were truncated at the 95th percentile to account for extreme values.^30^ Final analytic weights were created by multiplying the IPW by the NHATS survey weights, which accounted for the complex study design (i.e., stratum and cluster), attrition, and non-responses to the dried blood spot study. Second, we performed weighted ordinary least squares regression analyses. Two sets of models for each biomarker were presented. The first set of models applied the survey weights and adjusted for cohort membership. The second set of models applied the final analytic weights incorporating IPW and survey weights and adjusted for cohort membership. Statistical tests were two-sided, with a significance level of *p* < 0.05. Data analyses were performed using RStudio version 4.0.2,^31^ R package WeightIt,^32^ and STATA 17.^33^

We conducted several sensitivity analyses. Firstly, to validate the latent class group classification, we performed a group-based trajectory model and tested the agreement between the group membership from these two modeling strategies. Additionally, the replenished cohort was recruited later over the course of the survey with fewer observations. To test the reliability of the LCA results, we retrieved neighborhood physical disorder data for the replenished cohort from Round 7 to Round 11 so that both cohorts had an equal duration of observed repeated neighborhood physical disorder measures. We then re-estimated the latent class model based on this constructed data to assess agreement. Lastly, we tested alternative weight truncation at the 90th and 99th percentiles for the regression analyses.

## Results

Baseline descriptive statistics of the sample are presented in Table 1. The final analytical sample included 4,558 community-dwelling older adults, representing an estimated 36,502,275 U.S. older adults. In the weighted sample, 55% of the respondents were female, 80% were white, 8% were black, 7.5% were Hispanic, and other racial groups accounted for 4.5%. Older adults aged 65 to 74 accounted for 65% of the sample. Only 15.8% of the respondents had an education level lower than high school. 82% of these older adults lived in metropolitan areas. 7.5% reported currently experiencing financial strain.

**Table 1.** Descriptive Statistics of Study Sample at the Baseline.

| Variables (n, %) | Unweighted | With survey weights |
| --- | --- | --- |
| n | 4,558 | 36,502,275 |
| Female | 2,623 (57.5%) | 20,154,542 (55.2%) |
| Race |  |  |
| White | 3,296 (72.3%) | 29,244,403 (80.1%) |
| Black | 884 (19.4%) | 2,899,672 (7.9%) |
| Hispanic | 257 (5.6%) | 2,740,497 (7.5%) |
| Other | 121 (2.7%) | 1,617,702 (4.4%) |
| Education |  |  |
| Less than high school | 859 (18.8%) | 5,777,289 (15.8%) |
| High school | 1,529 (33.5%) | 11,875,356 (32.5%) |
| Some college | 907 (19.9%) | 7,773,732 (21.3%) |
| College and above | 1,263 (27.7%) | 11,075,897 (30.3%) |
| Age |  |  |
| 65 to 69 | 1,256 (27.6%) | 14,894,404 (40.8%) |
| 70 to 74 | 1,125 (24.7%) | 8,830,228 (24.2%) |
| 75 to 79 | 966 (21.2%) | 6,259,413 (17.1%) |
| 80 to 84 | 745 (16.3%) | 4,016,535 (11.0%) |
| 85 to 89 | 337 (7.4%) | 1,832,441 (5.0%) |
| 90+ | 129 (2.8%) | 669,253 (1.8%) |
| Marital status |  |  |
| Not married | 2,049 (45.0%) | 14,721,521 (40.3%) |
| Married or partnered | 2,509 (55.0%) | 21,780,754 (59.7%) |
| Current Smoking |  |  |
| No | 4,231 (92.8%) | 33,745,649 (92.4%) |
| Yes | 327 (7.2%) | 2,756,626 (7.6%) |
| Rented home | 678 (14.9%) | 5,285,915 (14.5%) |
| Childhood financial status |  |  |
| Poor | 1,688 (37.0%) | 12,564,865 (34.4%) |
| Average | 2,237 (49.1%) | 18,357,688 (50.3%) |
| Good | 633 (13.9%) | 5,579,721 (15.3%) |
| Born in the US | 4,148 (91%) | 32,366,358 (88.7%) |
| Having finance strain | 355 (7.8%) | 2,722,487 (7.5%) |
| Living in metropolitan | 3,626 (79.6%) | 29,780,433 (81.6%) |
| Cohort membership | 932 (20.4%) | 6,721,842 (18.4%) |
| 2011 cohort | 2,284 (50.1%) | 10,599,695 (29.0%) |
| 2015 cohort | 2,274 (49.9%) | 25,902,580 (71.0%) |
| Census region |  |  |
| Northeast Region: New England | 214 (4.7%) | 2,130,010 (5.8%) |
| Northeast Region: Middle Atlantic | 480 (10.5%) | 4,504,330 (12.3%) |
| Midwest Region: East North Central | 683 (15.0%) | 4,685,836 (12.8%) |
| Midwest Region: West North Central | 505 (11.1%) | 3,635,825 (10.0%) |
| South Region: South Atlantic | 1,003 (22.0%) | 7,563,260 (20.7%) |
| South Region: East South Central | 332 (7.3%) | 2,314,990 (6.3%) |
| South Region: West South Central | 511 (11.2%) | 4,191,965 (11.5%) |
| West Region: Mountain Division | 130 (2.9%) | 1,153,166 (3.2%) |
| West Region: Pacific Division | 700 (15.4%) | 6,322,892 (17.3%) |

*Note. Frequencies with percentages n (%) are presented.*

To identify patterns of neighborhood physical disorder, we performed latent class analyses and tested two- to five-group solutions. Model fit indices indicated that a four-class solution (BIC = 10485.29) yielded the best model fit (Supplementary Table S1). From this model, four distinct subgroups were identified (Figure 2). The majority of individuals in this sample exposed to low neighborhood physical disorder and this pattern is quite stable (3,889, 85.3%). In contrast, a small number of respondents had stable exposure to high neighborhood physical disorder (138, 3%). The remaining groups were characterized by either increased exposure (165, 4%) or decreased exposure (366, 8%) over time.

**Figure 2.**
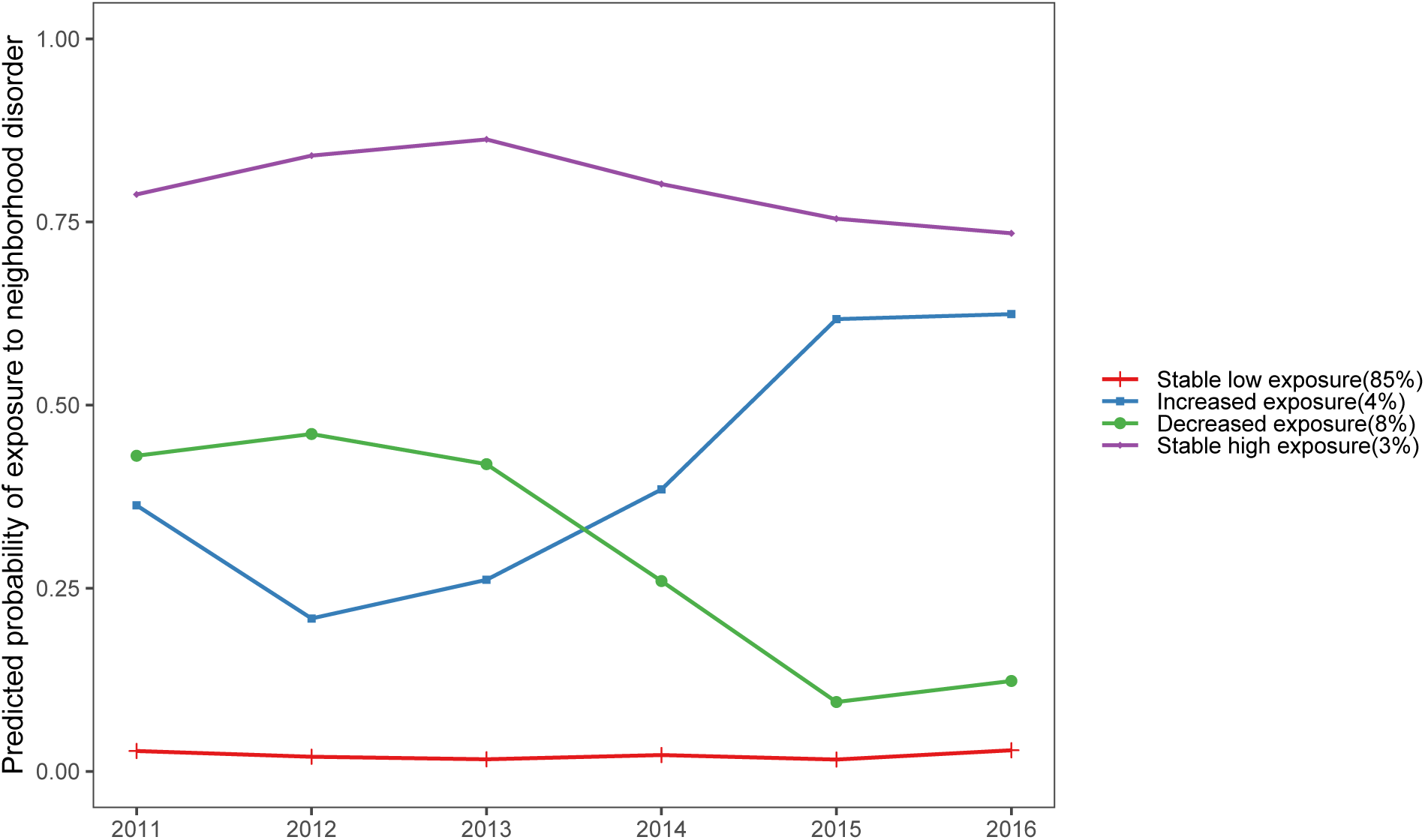
Predicted probability of exposure to neighborhood physical disorder from four-class classification over time (NHATS, 2011 - 2016)

Table 2 presents the characteristics of respondents across the four latent class subgroups. Older adults with stable exposure to high levels of disorder were more likely than those with constant low exposure, as well as more likely than other two groups, to be black (53%), Hispanic (21%), unmarried (70%), have less than high school education (53%), experience financial strain (26.8%), rent their homes (30%), currently smoke (21.7%), and be born outside the United States (17.4%). A similar pattern was observed among those living in neighborhoods with increased exposure or decreased exposure, compared to the stable low exposure group. Those with increased exposure were slightly worse off than those living in neighborhoods with decreased exposure.

**Table 2.** Baseline Sample Characteristics by Neighborhood Physical Disorder Subgroups.

| Variables (n, %) | Stable low exposure | Increased exposure | Decreased exposure | Stable high exposure | *p* |
| --- | --- | --- | --- | --- | --- |
| n | 3,889 (85.3%) | 165 (3.6%) | 366 (8.0%) | 138 (3.0%) |  |
| Female | 2,243 (57.7%) | 91 (55.2%) | 210 (57.4%) | 79 (57.2%) |  |
| Race |  |  |  |  |  |
| White | 3,044 (78.3%) | 54 (32.7%) | 165 (45.1%) | 33 (23.9%) | <0.001 |
| Black | 594 (15.3%) | 72 (43.6%) | 145 (39.6%) | 73 (52.9%) |  |
| Hispanic | 152 (3.9%) | 29 (17.6%) | 47 (12.8%) | 29 (21.0%) |  |
| Other | 99 (2.5%) | 10 (6.1%) | 9 (2.5%) | 3 (2.2%) |  |
| Education |  |  |  |  |  |
| Less than high school | 576 (14.8%) | 66 (40.0%) | 144 (39.3%) | 73 (52.9%) | <0.001 |
| High school | 1,313 (33.8%) | 55 (33.3%) | 114 (31.1%) | 47 (34.1%) |  |
| Some college | 802 (20.6%) | 26 (15.8%) | 63 (17.2%) | 16 (11.6%) |  |
| College and above | 1,198 (30.8%) | 18 (10.9%) | 45 (12.3%) | 2 (1.4%) |  |
| Age |  |  |  |  |  |
| 65 to 69 | 280 (7.2%) | 23 (13.9%) | 29 (7.9%) | 11 (8.0%) | 0.203 |
| 70 to 74 | 1,000 (25.7%) | 36 (21.8%) | 92 (25.1%) | 32 (23.2%) |  |
| 75 to 79 | 885 (22.8%) | 43 (26.1%) | 93 (25.4%) | 33 (23.9%) |  |
| 80 to 84 | 794 (20.4%) | 30 (18.2%) | 63 (17.2%) | 28 (20.3%) |  |
| 85 to 89 | 567 (14.6%) | 20 (12.1%) | 54 (14.8%) | 26 (18.8%) |  |
| 90+ | 363 (9.3%) | 13 (7.9%) | 35 (9.6%) | 8 (5.8%) |  |
| Marital status |  |  |  |  |  |
| Not Married | 1,628 (41.9%) | 106 (64.2%) | 219 (59.8%) | 96 (69.6%) | <0.001 |
| Married or partnered | 2,261 (58.1%) | 59 (35.8%) | 147 (40.2%) | 42 (30.4%) |  |
| Current Smoking |  |  |  |  |  |
| No | 3,652 (93.9%) | 148 (89.7%) | 323 (88.3%) | 108 (78.3%) | <0.001 |
| Yes | 237 (6.1%) | 17 (10.3%) | 43 (11.7%) | 30 (21.7%) |  |
| Rented house |  |  |  |  |  |
| No | 3,374 (86.8%) | 126 (76.4%) | 283 (77.3%) | 97 (70.3%) | <0.001 |
| Yes | 515 (13.2%) | 39 (23.6%) | 83 (22.7%) | 41 (29.7%) |  |
| Childhood financial status |  |  |  |  |  |
| Poor | 1,397 (35.9%) | 73 (44.2%) | 157 (42.9%) | 61 (44.2%) | 0.024 |
| Average | 1,941 (49.9%) | 74 (44.8%) | 163 (44.5%) | 59 (42.8%) |  |
| Good | 551 (14.2%) | 18 (10.9%) | 46 (12.6%) | 18 (13.0%) |  |
| Born in the US |  |  |  |  |  |
| No | 309 (7.9%) | 27 (16.4%) | 50 (13.7%) | 24 (17.4%) | <0.001 |
| Yes | 3,580 (92.1%) | 138 (83.6%) | 316 (86.3%) | 114 (82.6%) |  |
| Has finance strain |  |  |  |  |  |
| No | 3,648 (93.8%) | 141 (85.5%) | 313 (85.5%) | 101 (73.2%) | <0.001 |
| Yes | 241 (6.2%) | 24 (14.5%) | 53 (14.5%) | 37 (26.8%) |  |
| Urbanicity |  |  |  |  |  |
| Metropolitan | 3,092 (79.5%) | 128 (77.6%) | 293 (80.1%) | 113 (81.9%) | 0.821 |
| Non-metropolitan | 797 (20.5%) | 37 (22.4%) | 73 (19.9%) | 25 (18.1%) |  |
| Census region |  |  |  |  |  |
| Northeast Region: New England | 188 (4.8%) | 1 (0.6%) | 21 (5.7%) | 4 (2.9%) | <0.001 |
| Northeast Region: Middle Atlantic | 391 (10.1%) | 24 (14.5%) | 41 (11.2%) | 24 (17.4%) |  |
| Midwest Region: East North Central | 589 (15.1%) | 18 (10.9%) | 58 (15.8%) | 18 (13.0%) |  |
| Midwest Region: West North Central | 454 (11.7%) | 18 (10.9%) | 24 (6.6%) | 9 (6.5%) |  |
| South Region: South Atlantic | 830 (21.3%) | 46 (27.9%) | 82 (22.4%) | 45 (32.6%) |  |
| South Region: East South Central | 292 (7.5%) | 9 (5.5%) | 23 (6.3%) | 8 (5.8%) |  |
| South Region: West South Central | 406 (10.4%) | 27 (16.4%) | 57 (15.6%) | 21 (15.2%) |  |
| West Region: Mountain Division | 123 (3.2%) | 3 (1.8%) | 3 (0.8%) | 1 (0.7%) |  |
| West Region: Pacific Division | 616 (15.8%) | 19 (11.5%) | 57 (15.6%) | 8 (5.8%) |  |

Note. Frequencies with percentages *n* (%) are presented. $\chi^2$ tests were performed to compare group differences.

We further examined bivariate associations between the four LCA subgroups and the biomarkers (Supplementary Figure S1). Significant group differences were found in HbA1c, IL- 6, and hsCRP. Older adults who were exposed to stable high neighborhood physical disorder showed the highest levels of IL-6 and hsCRP. Those with stable high or increased exposure had higher levels of HbA1c compared to the other two groups.

Next, we conducted IPW regression analyses. Applying IPW successfully balanced all covariates across neighborhood physical disorder subgroups. Supplementary Figure S2 shows that standardized differences for all covariates were less than 0.1 and the Kolmogorov-Smirnov statistics were non-significant after applying IPW. Findings from the models with and without IPW adjustment were presented in Figure 3 and Supplementary Table S2. In the IPW weighted models, older adults with stable high exposure reported higher BMI (*b* = 0.06, 95% CI: 0.01- 0.11). Those with increased exposure (*b* = 0.05, 95% CI: 0.02-0.08), decreased exposure (*b* = 0.03, 95% CI: 0.01-0.06), and stable high exposure (*b* = 0.09, 95% CI: 0.05-0.13) to neighborhood physical disorder showed significantly higher levels of HbA1c, compared to those with stable low exposure. Additionally, older adults living in neighborhoods with decreased disorder and stable high disorder had significantly higher levels of inflammation, as indicated by higher levels of IL-6 (*b* = 0.18, 95% CI: 0.01-0.36; *b* = 0.21, 95% CI: 0.04-0.36), and hsCRP (*b* = 0.12, 95% CI: 0.01-0.22; *b* = 0.22, 95% CI: 0.03-0.41), compared to their counterparts with stable low exposure. The results from models that applied survey weights without IPW were generally consistent with the IPW adjusted results.

**Figure 3.**
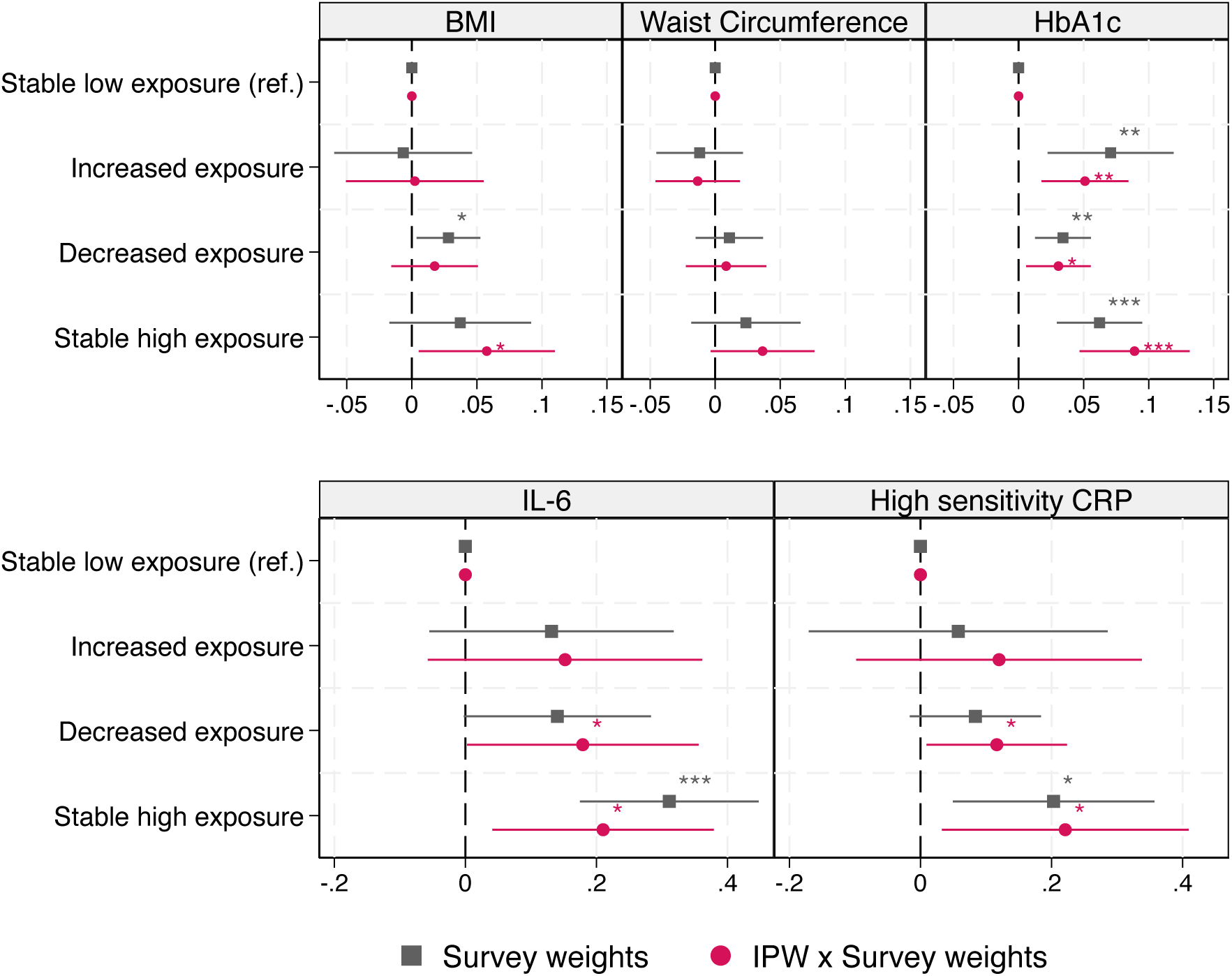
Regression coefficients of metabolic and inflammatory biomarkers (log transformed) and neighborhood physical disorder subgroups (NHATS, 2011-2017) *Note:* 1. Stable low exposure group was the reference group. All models adjusted for the binary indicator of cohort membership. 2. Two sets of regression coefficients are presented. The first set of models applied survey weights and the second set of models applied analytic weights incorporating IPW and survey weights. BMI: body mass index; HbA1c: hemoglobin A1C; IL-6: interleukin-6; CRP: C-reactive protein. *\* p*<0.05, *\*\* p*<0.01, *\*\*\* p*<0.001.

In the sensitivity analyses (Supplementary Figure S3), we found substantial agreement between our current group classification and when applying group-based trajectory models (kappa = 0.6) as well as using the constructed dataset (kappa = 0.7). This indicates that our LCA results were robust under various models and data setups. Inferences remained unchanged in sensitivity analyses using alternative weight truncation (results not shown).

## Discussion

This study examined the association between risk patterns of exposure to neighborhood physical disorder and metabolic and inflammatory biomarkers among a nationally representative sample of older adults. We identified four subgroups representing various trajectories of stability and changes in neighborhood contexts. Results from the LCA revealed that approximately 85% of respondents lived in neighborhoods featured by stable low physical disorder, while a small portion (3%) experienced stable exposure to high disorder, suggesting that most respondents had stable neighborhood trajectories. This finding is consistent with previous research showing little or no change in neighborhood contexts among community-dwelling older people. ^22,34^ Notably, racial/ethnic minority groups, individuals with lower education, and those with financial strains were disproportionately represented in the stable high exposure group, suggesting the persistent residential stratification along racial, ethnic, and socioeconomic lines. ^24,35^

Living in neighborhoods with stable high physical disorder is associated with higher subsequent levels of metabolic and inflammatory biomarkers, including IL-6, hsCRP, and HbA1c. These associations remained statistically significant after accounting for individual-level characteristics that could affect both exposure to neighborhood physical disorder and biomarker levels. Specifically, we found that older adults with stable high exposure to neighborhood physical disorder had elevated levels of IL-6 and hsCRP. This relationship is consistent with clinical evidence suggesting that the inflammatory response is a crucial pathway linking neighborhood deprivation to health outcomes among community-dwelling older adults. ^6,13,18,20^

Our research revealed pronounced associations between neighborhood physical disorder and HbA1c, while prior clinical and observational studies have reported weak or non-significant associations. ^36,37^ This discrepancy may be attributed to variations in sample size and neighborhood measurement methods. Our study used a nationally representative sample and standardized observer assessments of neighborhoods, in contrast to studies that often relied on limited regional samples or participants’ subjective perceptions of their residential environments. Our results aligned with previous studies showing the detrimental effect of cumulative exposure to neighborhood physical disorder on HbA1c in the NHATS sample. ^13^ This finding suggests the potential long-term consequences of residing in disadvantaged neighborhoods on metabolic health.

While neighborhood physical disorder is associated with metabolic and inflammatory biomarkers, the relationships with anthropometric adiposity measures, such as BMI and waist circumference, are weak. Although evidence from systematic reviews confirmed a general link between neighborhood socioeconomic status and BMI, ^19,38^ findings regarding neighborhood disorder and BMI are mixed. Similar to our findings, Letarte and colleagues (2022) found that residing in neighborhoods characterized by deprived upward, deprived downward, and stable deprived trajectories was significantly associated with obesity compared to those living in stable low deprived neighborhoods. Moreover, substantial heterogeneities also exist in neighborhood effects on weight status. For instance, using a sample from the Baltimore Memory Study, Glass and colleagues (2006) found non-significant associations between neighborhood deprivation and obesity for both Black and White older adults.^39^ However, Keita et al. (2014) reported that living in a deprived neighborhood was associated with higher BMI and larger waist circumference among White middle-aged and older adults, but not among their Black counterparts. Some research also suggests stronger neighborhood effects on women than on men. ^11,40^ Due to the small sample size in the stable high exposure group, we were not able to conduct analyses examining gender or racial/ethnic differences. Future studies are needed to examine the mechanisms underlying the heterogeneity of neighborhood effects across social-demographic groups.

Our findings have important implications. In clinical settings, social determinants of health should be incorporated into patient care. Healthcare practitioners should consider screening patients for stressors related to environmental risks and neighborhood hazards. Identifying patients living in high-disorder neighborhoods can help clinicians recognize those at greater risk for various health conditions. Furthermore, practitioners may not be able to directly address issues like neighborhood physical disorder, their referrals to social services and community resources can be crucial for patients affected by neighborhood physical disorder. This may include connecting them with programs that offer mental health support, housing assistance, or community health initiatives aimed at improving living conditions.^13^ While our study highlights the association between neighborhood physical disorder and biological risk, recent randomized controlled trials provide causal evidence supporting the positive influences of neighborhood interventions on health outcomes. For instance, a citywide cluster randomized controlled trial in Philadelphia demonstrated that greening vacant lots significantly reduced stress and improved mental health among residents. ^41^ This evidence suggests that addressing neighborhood physical disorder can be effective through community-level interventions and programs. Policy efforts focusing on investing in safer streets, green spaces, and recreational facilities can promote physical activity and reduce stress.^42^ To better address issues like littering and graffiti, policies could involve increasing community-based cleanup initiatives, installing more public trash receptacles, and creating designated spaces for street art for the reduction of environmental risks and the promotion of healthy aging in place. ^13^

There are several limitations of this study. First, the measurement of neighborhood physical disorder in the NHATS survey includes only three items, which may not capture other important forms of neighborhood physical disorder, such as noise. It is possible that we underestimated the prevalence of neighborhood physical disorder in this sample. Second, despite using IPW to adjust for confounders, residual confounding might persist, especially if unmeasured confounders influenced both neighborhood physical disorder and health outcomes. Third, the small sample sizes of the LCA subgroups—particularly those living in neighborhoods with increased exposure, decreased exposure, and stable high exposure —may limit the statistical power of the analysis, which could lead to an underestimation of the effects of neighborhood physical disorder. Further studies with larger longitudinal samples are required to better validate the results from those groups. Lastly, we included six rounds of neighborhood measures to examine exposure to neighborhood physical disorder over time. However, six years may not be long enough to capture the time frame relevant to the outcomes we studied. Additional studies with longer follow-ups are needed to disentangle the cumulative effects of neighborhood physical disorder and strengthen causal inference of neighborhood contexts on health.

This study has several important strengths. The study was conducted using a nationally representative sample to strengthen the generalizability of the results. Additionally, we leveraged the longitudinal design of the NHATS data to examine exposure histories to neighborhood physical disorder, which allowed us to assess the dynamic changes of neighborhoods that cross-sectional measures are unable to capture. Moreover, we implemented a machine learning based IPW approach to mimic a quasi-experimental design and balance individual characteristics across different neighborhood physical disorder subgroups, thereby mitigating potential confounding effects. Lastly, while previous studies found perceived neighborhood physical disorder being significantly associated with various health outcomes,^10^ using objective rather than perceived neighborhood physical disorder measures likely reduced recall bias and social desirability bias, therefore enhancing the validity of our findings.

In conclusion, our findings reveal significant associations between long-term exposure to neighborhood physical disorder and metabolic and inflammatory biomarkers among older adults. Specifically, those with stable high exposure to neighborhood physical disorder exhibit higher levels of metabolic and inflammatory biomarkers compared to their counterparts with stable low exposure. These results underscore the critical role that immediate residential environments play in influencing physiological aging processes. Incorporating screenings for social determinants of health into routine healthcare visits for older adults could be a viable way to improve health outcomes and supporting healthy aging in place. Additionally, addressing neighborhood physical disorder through targeted policies and community initiatives can be pivotal in mitigating environmental health risks.

## Conflict of Interest

The authors declare that there is no conflict of interest.

## Funding

The preparation of this article was supported in part by the National Institute on Aging of the National Institutes of Health (R01AG077529, P30AG021342) and the University of Minnesota Life Course Center on the Demography and Economics of Aging (P30AG066613), funded through a grant from the National Institute on Aging. The National Health and Aging Trends Study is sponsored by the National Institute on Aging (U01AG032947) and was conducted by Johns Hopkins University and the University of Michigan. The content is solely the responsibility of the authors and does not necessarily represent the official views of the National Institutes of Health.

## Data Availability

The National Health and Aging Trends Study can be accessed via https://nhats.org/researcher. Statistical codes for this study are made available by email request to the corresponding authors.

## Acknowledgements

The author’s contributions were as follows: J.Y. contributed to the study design, funding acquisition, data analyses, and drafting the manuscript; T. KM. C, and W. S. M. provided feedback and revised the manuscript. X.C. contributed to funding acquisition, reviewing, and editing the manuscript.

---

**Supplementary Figure S1.**
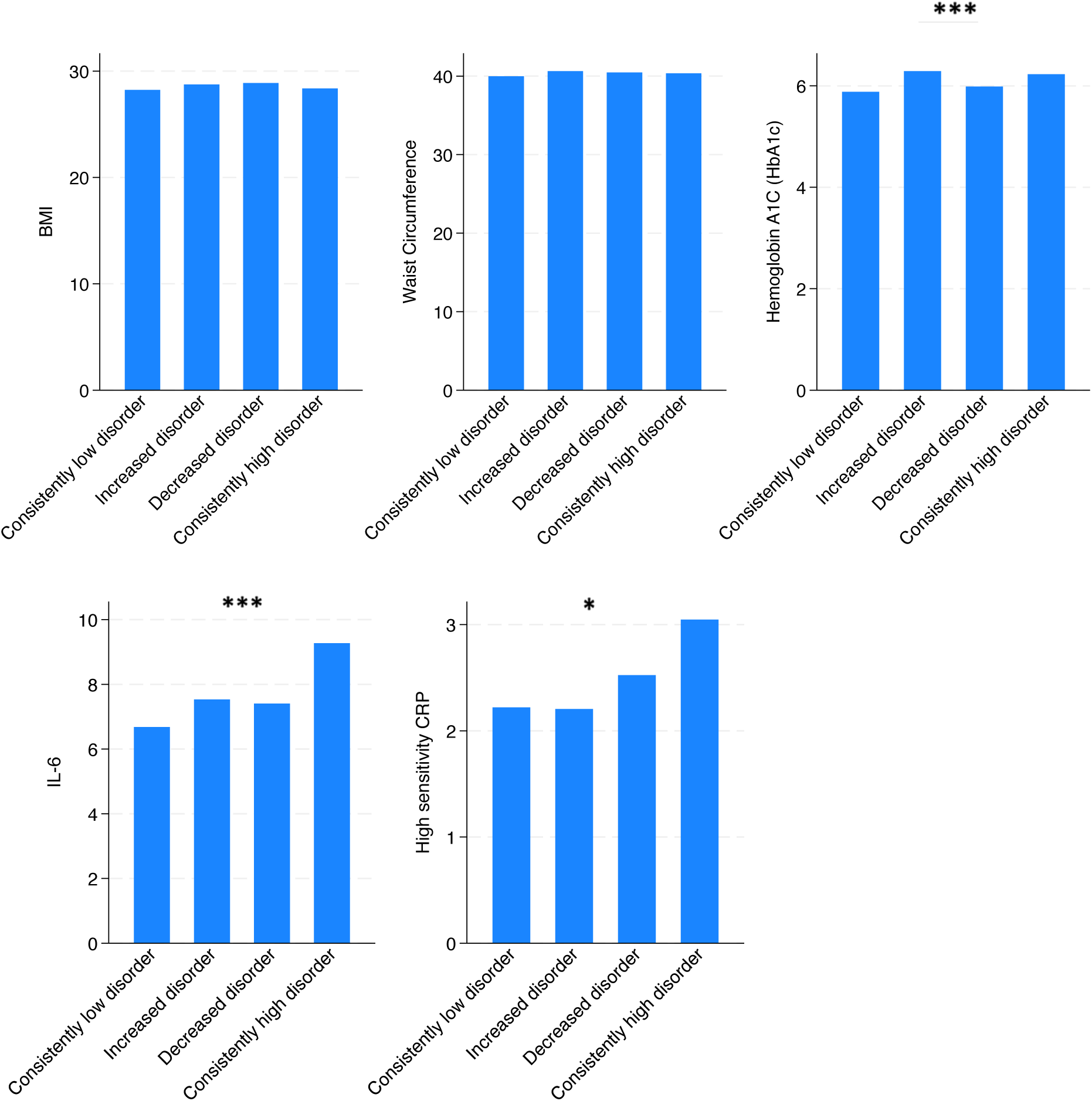
Bivariate distribution of metabolic and inflammatory biomarkers across neighborhood physical disorder groups (with survey weights) Note: Kruskal-Wallis rank sum tests were performed to compare statistical differences across groups. BMI: body mass index; CRP: High-sensitivity C-reactive protein; IL-6: interleukin-6. Survey weights were applied to allow inferences to be drawn to US older adult Medicare beneficiaries. *\*p < .05, **p < .01, ***p < .001*.

**Supplementary Figure S2.**
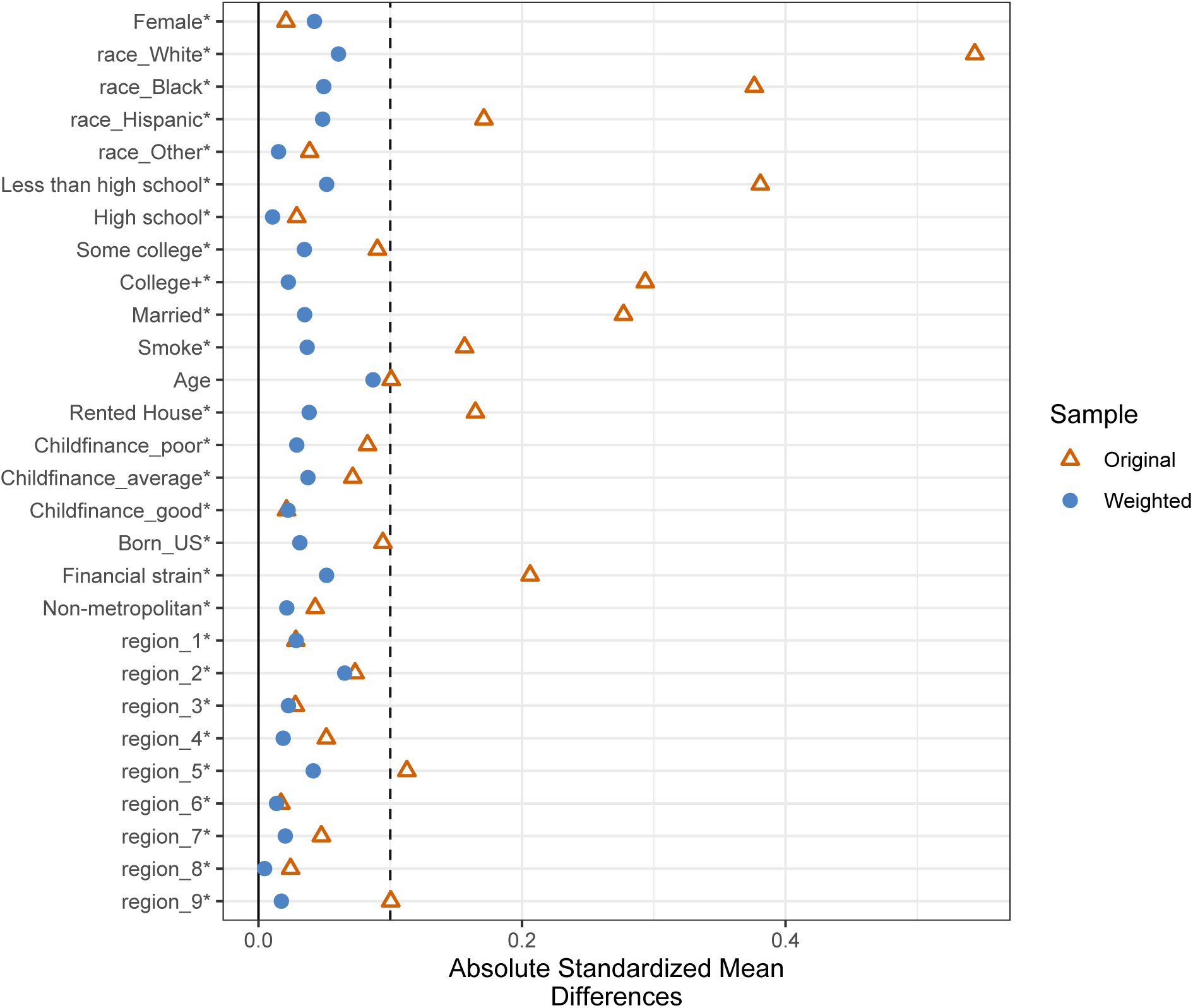
Covariates balance plot before and after inverse probability weighting Note: After applying IPWs, all covariates had a standardized difference of less than 0.1 and the Kolmogorov-Smirnov statistic was non-significant. All covariates were balanced after applying the GBM weighting adjustment. Region_1: Northeast Region: New England; Region_2: Northeast Region: Middle Atlantic; Region_3: Midwest Region: East North Central; Region_4: Midwest Region: West North Central; Region_5: South Region: South Atlantic; Region_6: South Region: East South Central; Region_7: South Region: West South Central; Region_8: West Region: Mountain Division; Region_9: West Region: Pacific Division.

**Supplementary Figure S3.**
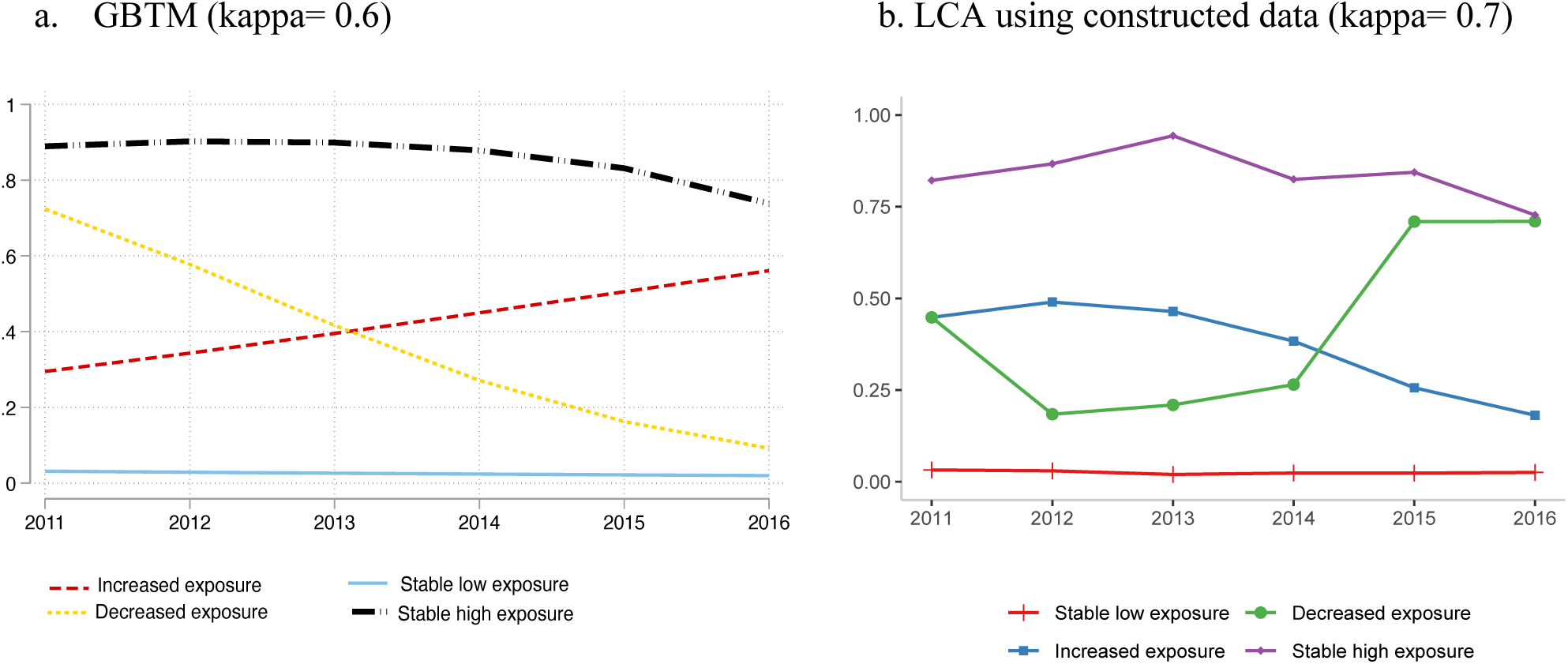
Sensitivity Analysis of Latent Class Results Note. Fig. S3a (left panel) presents neighborhood physical disorder categorization using group-based trajectory modeling (GBTM). Fig. S3b (right panel) presents Latent Class Analysis (LCA) findings using a constructed dataset so that two cohorts had an equal duration of repeated neighborhood disorder measures. Cohen’s Kappa coefficient indicates the substantial agreement of GBTM grouping with the grouping presented in the main analysis (Fig. S3a: κ=0.6). The LCA grouping results using constructive data also show substantial agreement with the grouping presented in the main analysis (Fig. S3b: κ=0.7). Kappa coefficient ranges from -1 to 1, with higher values indicating higher agreement. Specifically, κ>0.6 indicate substantial agreement. κ>0.8 indicate almost perfect agreement.

**Supplementary Table S1.** Latent Class Analysis Model Fit Statistics.

|  | Log Likelihood | Degree of<br>freedom | BIC |
| --- | --- | --- | --- |
| Class 2 | -5190.227 | 13 | 10489.95 |
| Class 3 | -5159.636 | 20 | 10487.73 |
| Class 4 | -5128.939 | 27 | 10485.29 |
| Class 5 | -5117.957 | 34 | 10522.28 |

Note: BIC=Bayesian information criterion; BIC is used to assess goodness of fit with smaller values indicating better fit.

**Supplementary Table S2.** Regression coefficients of metabolic and inflammatory biomarkers and four class neighborhood physical disorder.

|  | Model 1 | Model 2 | Model 3 | Model 4 | Model 5 |
| --- | --- | --- | --- | --- | --- |
|  | BMI | Waist<br>Circumference | HbA1c | IL-6 | hsCRP |
| **Panel 1: Regression with analytic weights incorporating IPW and survey weights** |  |  |  |  |  |
| Increased exposure | 0.002 | -0.01 | 0.05** | 0.15 | 0.12 |
|  | [-0.05, 0.06] | [-0.05, 0.02] | [0.02, 0.08] | [-0.06, 0.36] | [-0.10, 0.34] |
| Decreased exposure | 0.02 | 0.01 | 0.03* | 0.18* | 0.12* |
|  | [-0.02, 0.05] | [-0.02, 0.04] | [0.01, 0.06] | [0.01, 0.36] | [0.01, 0.22] |
| High stable exposure | 0.06* | 0.04 | 0.09*** | 0.21* | 0.22* |
|  | [0.01, 0.11] | [-0.00, 0.08] | [0.05, 0.13] | [0.04, 0.38] | [0.03, 0.41] |
| Intercept | 3.33** | 3.70** | 1.92** | 1.73** | 0.94** |
|  | [3.32, 3.34] | [3.69, 3.71] | [1.92, 1.93] | [1.69, 1.77] | [0.90, 0.99] |
| **Panel 2: Regression with survey weights** |  |  |  |  |  |
| Increased exposure | -0.01 | -0.01 | 0.07** | 0.13 | 0.06 |
|  | [-0.06, 0.05] | [-0.05, 0.02] | [0.02, 0.12] | [-0.05, 0.32] | [-0.17, 0.29] |
| Decreased exposure | 0.03* | 0.01 | 0.03** | 0.14 | 0.08 |
|  | [0.00, 0.05] | [-0.01, 0.04] | [0.01, 0.06] | [-0.00, 0.28] | [-0.02, 0.18] |
| High stable exposure | 0.04 | 0.02 | 0.06*** | 0.31*** | 0.20* |
|  | [-0.02, 0.09] | [-0.02, 0.07] | [0.03, 0.10] | [0.17, 0.45] | [0.05, 0.36] |
| Intercept | 3.34** | 3.70** | 1.93** | 1.74** | 0.94** |
|  | [3.33, 3.35] | [3.69, 3.71] | [1.92, 1.93] | [1.70, 1.78] | [0.91, 0.98] |

*Note:* Coefficients and 95% confidence intervals were reported. Biomarkers were log transformed.
The low stable exposure group was the reference group. All regression models adjusted for the binary indicator of cohort membership.
BMI: body mass index; HbA1c: hemoglobin A1C; IL-6: interleukin-6; HsCRP: High-sensitivity C-reactive protein.
\* $p < 0.05$ , \*\* $p < 0.01$ , \*\*\* $p < 0.001$ .

## Notes

### Competing Interest Statement

The authors have declared no competing interest.

### Author Declarations

The study used publicly available data from the National Health and Aging Trends Study. The National Health and Aging Trends Study can be accessed via https://nhats.org/researcher.

## References

1. Sampson RJ, Raudenbush SW. Seeing Disorder: Neighborhood Stigma and the Social Construction of “Broken Windows.” Soc Psychol Q. 2004;67(4):319–342. doi:10.1177/019027250406700401

2. Glymour MM, Mujahid M, Wu Q, White K, Tchetgen Tchetgen EJ. Neighborhood Disadvantage and Self-Assessed Health, Disability, and Depressive Symptoms: Longitudinal Results From the Health and Retirement Study. Annals of Epidemiology. 2010;20(11):856–861. doi:10.1016/j.annepidem.2010.08.003

3. Gustafsson PE, San Sebastian M, Janlert U, Theorell T, Westerlund H, Hammarström A. Life-Course Accumulation of Neighborhood Disadvantage and Allostatic Load: Empirical Integration of Three Social Determinants of Health Frameworks. Am J Public Health. 2014;104(5):904–910. doi:10.2105/AJPH.2013.301707

4. Keysor JJ, Jette AM, LaValley MP, et al. Community Environmental Factors Are Associated With Disability in Older Adults With Functional Limitations: The MOST Study. The Journals of Gerontology: Series A. 2010;65A(4):393–399. doi:10.1093/gerona/glp182

5. Wang F, Qin W, Yu J. Neighborhood Social Cohesion and Mobility Limitations Among Community-dwelling Older Americans: The Mediating Roles of Depressive Symptoms and Mastery. Int J Aging Hum Dev. 2022;94(3):290–311. doi:10.1177/00914150211037657

6. Roberts LC, Schwartz BS, Samuel LJ. Neighborhood Characteristics and Cardiovascular Biomarkers in Middle-Aged and Older Adults: the Baltimore Memory Study. J Urban Health. 2021;98(1):130–142. doi:10.1007/s11524-020-00499-7

7. Selvin E, Steffes MW, Zhu H, et al. Glycated Hemoglobin, Diabetes, and Cardiovascular Risk in Nondiabetic Adults. New England Journal of Medicine. 2010;362(9):800–811. doi:10.1056/NEJMoa0908359

8. Black PH. The inflammatory response is an integral part of the stress response: Implications for atherosclerosis, insulin resistance, type II diabetes and metabolic syndrome X. *Brain*, Behavior, and Immunity. 2003;17(5):350–364. doi:10.1016/S0889-1591(03)00048-5

9. Emerging Risk Factors Collaboration, Kaptoge S, Di Angelantonio E, et al. C-reactive protein concentration and risk of coronary heart disease, stroke, and mortality: an individual participant meta-analysis. Lancet. 2010;375(9709):132–140. doi:10.1016/S0140-6736(09)61717-7

10. Nguyen AW, Taylor HO, Lincoln KD, et al. Neighborhood Characteristics and Inflammation Among Older Black Americans: The Moderating Effects of Hopelessness and Pessimism. The Journals of Gerontology: Series A. 2022;77(2):315–322. doi:10.1093/gerona/glab121

11. Robert SA, Reither EN. A multilevel analysis of race, community disadvantage, and body mass index among adults in the US. Social Science & Medicine. 2004;59(12):2421–2434. doi:10.1016/j.socscimed.2004.03.034

12. Letarte L, Samadoulougou S, McKay R, Quesnel-Vallée A, Waygood EOD, Lebel A. Neighborhood deprivation and obesity: Sex-specific effects of cross-sectional, cumulative and residential trajectory indicators. Social Science & Medicine. 2022;306:115049. doi:10.1016/j.socscimed.2022.115049

13. Roberts Lavigne LC, Tian J, Hladek M, LaFave SE, Szanton SL, Samuel LJ. Residential Street Block Disorder and Biological Markers of Aging in Older Adults: The National Health and Aging Trends Study. The Journals of Gerontology: Series A. 2021;76(11):1969–1976. doi:10.1093/gerona/glab166

14. Buschmann RN, Prochaska JD, Cutchin MP, Peek MK. Stress and health behaviors as potential mediators of the relationship between neighborhood quality and allostatic load. Annals of Epidemiology. 2018;28(6):356–361. doi:10.1016/j.annepidem.2018.03.014

15. Brisson D, McCune S, Wilson JH, Speer SR, McCrae JS, Hoops Calhoun K. A Systematic Review of the Association between Poverty and Biomarkers of Toxic Stress. Journal of Evidence-Based Social Work. 2020;17(6):696–713. doi:10.1080/26408066.2020.1769786

16. Muñoz E, Scott SB, Corley R, Wadsworth SJ, Sliwinski MJ, Reynolds CA. The role of neighborhood stressors on cognitive function: A coordinated analysis. Health & Place. 2020;66:102442. doi:10.1016/j.healthplace.2020.102442

17. Juster RP, McEwen BS, Lupien SJ. Allostatic load biomarkers of chronic stress and impact on health and cognition. Neuroscience & Biobehavioral Reviews. 2010;35(1):2–16. doi:10.1016/j.neubiorev.2009.10.002

18. Petersen KL, Marsland AL, Flory J, Votruba-Drzal E, Muldoon MF, Manuck SB. Community Socioeconomic Status is Associated With Circulating Interleukin-6 and C-Reactive Protein. Psychosomatic Medicine. 2008;70(6):646. doi:10.1097/PSY.0b013e31817b8ee4

19. Mohammed SH, Habtewold TD, Birhanu MM, et al. Neighbourhood socioeconomic status and overweight/obesity: a systematic review and meta-analysis of epidemiological studies. BMJ Open. 2019;9(11):e028238. doi:10.1136/bmjopen-2018-028238

20. Keita AD, Judd SE, Howard VJ, Carson AP, Ard JD, Fernandez JR. Associations of neighborhood area level deprivation with the metabolic syndrome and inflammation among middle- and older- age adults. BMC Public Health. 2014;14(1):1319. doi:10.1186/1471-2458-14-1319

21. Murray ET, Diez Roux AV, Carnethon M, Lutsey PL, Ni H, O’Meara ES. Trajectories of Neighborhood Poverty and Associations With Subclinical Atherosclerosis and Associated Risk Factors. Am J Epidemiol. 2010;171(10):1099–1108. doi:10.1093/aje/kwq044

22. Jivraj S, Murray ET, Norman P, Nicholas O. The impact of life course exposures to neighbourhood deprivation on health and well-being: a review of the long-term neighbourhood effects literature. Eur J Public Health. 2019;30(5):922–928. doi:10.1093/eurpub/ckz153

23. Oakes JM, Andrade KE, Biyoow IM, Cowan LT. Twenty Years of Neighborhood Effect Research: An Assessment. Curr Epidemiol Rep. 2015;2(1):80–87. doi:10.1007/s40471-015-0035-7

24. Osypuk TL, Acevedo-Garcia D. Beyond individual neighborhoods: A geography of opportunity perspective for understanding racial/ethnic health disparities. Health & Place. 2010;16(6):1113–1123. doi:10.1016/j.healthplace.2010.07.002

25. Freedman VA, Kasper JD. Cohort Profile: The National Health and Aging Trends Study (NHATS). International Journal of Epidemiology. 2019;48(4):1044–1045g. doi:10.1093/ije/dyz109

26. Kasper J, Skehan M, Seeman T, Freedman V. Dried Blood Spot (DBS) Based Biomarkers in the National Health and Aging Trends Study User Guide: Final Release. Baltimore: Johns Hopkins University Bloomberg School of Public Health. Published online 2019.

27. Samuel LJ, Hladek M, Tian J, Roberts Lavigne LC, LaFave SE, Szanton SL. Propensity score weighted associations between financial strain and subsequent inflammatory biomarkers of aging among a representative sample of U.S. older adults. BMC Geriatrics. 2022;22(1):467. doi:10.1186/s12877-022-03112-5

28. Yu J, Qin W, Huang W, Thomas K. Oral Health and Mortality Among Older Adults: A Doubly Robust Survival Analysis. American Journal of Preventive Medicine. 2023;64(1):9–16. doi:10.1016/j.amepre.2022.08.006

29. McCaffrey DF, Ridgeway G, Morral AR. Propensity score estimation with boosted regression for evaluating causal effects in observational studies. Psychol Methods. 2004;9(4):403–425. doi:10.1037/1082-989X.9.4.403

30. Austin PC, Stuart EA. Moving towards best practice when using inverse probability of treatment weighting (IPTW) using the propensity score to estimate causal treatment effects in observational studies. Statistics in Medicine. 2015;34(28):3661–3679. doi:10.1002/sim.6607

31. R Core Team. R: A Language and Environment for Statistical Computing. R Foundation for Statistical Computing; 2021. https://www.R-project.org/

32. Greifer N. WeightIt: Weighting for Covariate Balance in Observational Studies. Published online 2023. https://ngreifer.github.io/WeightIt/, https://github.com/ngreifer/WeightIt

33. StataCorp. Stata Statistical Software: Release 17. College Station, TX: StataCorp LLC.; 2021.

34. Gill TM, Becher RD, Leo-Summers L, Gahbauer EA. Changes in neighborhood disadvantage over the course of 22 years among community-living older persons. Journal of the American Geriatrics Society. n/a(n/a). doi:10.1111/jgs.19172

35. Acevedo-Garcia D, Lochner KA, Osypuk TL, Subramanian SV. Future Directions in Residential Segregation and Health Research: A Multilevel Approach. Am J Public Health. 2003;93(2):215–221. doi:10.2105/AJPH.93.2.215

36. Gary TL, Safford MM, Gerzoff RB, et al. Perception of Neighborhood Problems, Health Behaviors, and Diabetes Outcomes Among Adults With Diabetes in Managed Care: The Translating Research Into Action for Diabetes (TRIAD) Study. Diabetes Care. 2008;31(2):273–278. doi:10.2337/dc07-1111

37. Walker RJ, Garacci E, Palatnik A, Ozieh MN, Egede LE. The Longitudinal Influence of Social Determinants of Health on Glycemic Control in Elderly Adults With Diabetes. Diabetes Care. 2020;43(4):759–766. doi:10.2337/dc19-1586

38. Mackenbach JD, Rutter H, Compernolle S, et al. Obesogenic environments: a systematic review of the association between the physical environment and adult weight status, the SPOTLIGHT project. BMC Public Health. 2014;14(1):233. doi:10.1186/1471-2458-14-233

39. Glass TA, Rasmussen MD, Schwartz BS. Neighborhoods and Obesity in Older Adults: The Baltimore Memory Study. American Journal of Preventive Medicine. 2006;31(6):455–463. doi:10.1016/j.amepre.2006.07.028

40. Wen M, Maloney TN. Latino Residential Isolation and the Risk of Obesity in Utah: The Role of Neighborhood Socioeconomic, Built-Environmental, and Subcultural Context. J Immigrant Minority Health. 2011;13(6):1134–1141. doi:10.1007/s10903-011-9439-8

41. South EC, Hohl BC, Kondo MC, MacDonald JM, Branas CC. Effect of Greening Vacant Land on Mental Health of Community-Dwelling Adults: A Cluster Randomized Trial. JAMA Network Open. 2018;1(3):e180298. doi:10.1001/jamanetworkopen.2018.0298

42. Carpenter M. From ‘healthful exercise’ to ‘nature on prescription’: The politics of urban green spaces and walking for health. Landscape and Urban Planning. 2013;118:120–127. doi:10.1016/j.landurbplan.2013.02.009

